# *OXTR rs53576* variation with breast and nipple pain in breastfeeding women

**DOI:** 10.1101/2020.09.21.20199208

**Authors:** Ruth Lucas, Yiming Zhang, Stephen J. Walsh, Angela Starkweather, Erin Young

## Abstract

**Purpose:** To evaluate associations among breast and nipple pain sensitivity and candidate pain sensitivity single-nucleotide polymorphisms [SNPs], (*COMT* rs6269, rs4633, rs4818, rs4680 and *OXTR* rs2254298, rs53576) in breastfeeding women.

**Design:** A secondary analysis of sixty women participating in a pilot randomized controlled trial of a pain self-management intervention.

**Methods:** All participants underwent standardized mechanical somatosensory testing for an assessment of pain sensitivity and provided baseline buccal swabs for genetic analysis. At 1, 2, and 6 weeks postpartum, women self-reported breast and nipple pain severity using a visual analogue scale.

**Results:** Women with the minor allele *OXTR* rs53576 reported 8.18-fold higher breast and nipple pain severity over time. For every 1-unit increase in mechanical detection threshold and windup ratio, women reported 16.51-fold and 4.82-fold higher breast and nipple pain severity respectively. Six women with the *OXTR* rs2254298 minor allele reported allodynia.

**Discussion:** The presence of *OXTR* alleles in women with enhanced pain sensitivity suggests a phenotype of genetic risk for ongoing breast and nipple with potential for pain-associated breastfeeding cessation. Somatosensory testing identified women who reported higher breast and nipple pain during the first weeks of breastfeeding.

**Clinical Implications:** Pain sensitivity testing can help to identify women at risk of intolerable and/or ongoing breastfeeding pain who may benefit from additional support to mitigate early breastfeeding cessation. Targeted interventions are needed to address breastfeeding pain, including management of infant latch, positioning, and infection as well as support for self-management of breastfeeding pain.

## Background

Most women report breast and nipple discomfort in the first weeks of initiating breastfeeding (Declercq, Sakala, Corry, Applebaum, Herrlich, 2013; Lucas & McGrath, 2016). However, 30% of new mothers will cease breastfeeding in the first 3 weeks due to intolerable pain (Odom, Li, Scanlon, Perrine, & Grummer-Strawn, 2013). Among women who seek professional lactation support during the first 30 days of breastfeeding, 30% of women require assistance with ongoing breast and nipple pain and ≤50% report resolution of their pain (Kent et al., 2015). Thus, intolerable and/or ongoing breast and nipple pain during breastfeeding is a clinically significant issue and a major barrier for continued breastfeeding. It is unknown if there is a molecular risk for intolerable and/or ongoing breast and nipple pain during breastfeeding that may influence the likelihood of breastfeeding cessation.

Breastfeeding is the most effective preventative measure for protecting infant health, supports mother and infant emotional communication, and provides health promotion and protection to women and infants across the lifespan (Quigley, Carson, Sacker, & Kelly, 2016; Victora et al., 2016). In 2018, 83.2% women initiated breastfeeding, but within 2-3 weeks after delivery the rate of exclusive breastfeeding dropped to 60% and 53% by 2 months as women returned to work (Center for Disease Control and Prevention, 2018). However, in order to have adequate milk supply for mothers and infants to breastfeed to 6 months, the public health goal for optimal health benefit for women and infants, women need to routinely breastfeed or pump for at least 9 weeks (Dozier et al., 2018). Although the reasons for early breastfeeding cessation can be complex, including psychological and social determinants of health, women most often report ceasing breastfeeding due to the lack of infant satiation, perception of maternal milk insufficiency, and intolerable and/or ongoing breast and nipple pain (Center for Disease Control and Prevention, 2018).

Pain during breastfeeding arises from complex stimulation of the glandular, somatic, and visceral tissues which are transmitted via nociceptors pathways that can become sensitized leading to altered sensation such as allodynia or spontaneous pain (Eriksson, Lindh, Uvnäs-Moberg, & Hökfelt, 1996; Jackson, Mandler, & O’Keefe-McCarthy, 2019). Biologic factors (infection) and anatomic factors (positioning challenges, infant anatomic issue such as ankyloglossia or torticollis, abnormal nipple shape or size) are among the triggers for sensitization of nociceptor pathways and ongoing pain (Berens, Eglash, Malloy, & Steube, 2016; Lucas & McGrath, 2016). In addition, breastfeeding occurs during a period of intense emotional and neurohormonal changes known to impact an individuals’ pain sensitivity (Amir, Jones, & Buck, 2015; Office of the Assistant Secretary for Health, 2019; Schug et al., 2019). Emotional reactions to pain can suppress or amplify the perception of painful stimuli (Edwards, Campbell, Jamison, & Wiech, 2009; Russell, Lincoln, & Starkweather, 2018). All of these factors together may contribute to ongoing pain which interferes with milk ejection, nipple expansion, and creates a negative neurohormonal feedback loop leading to maternal milk insufficiency and a lack of infant satiation (Eriksson et al., 1996; Francis & Dickton, 2019; Newton & Newton, 1948). However, less studied has been the role of individual variation in pain sensitivity and its influence on breastfeeding pain and duration.

Experimental pain methods using quantitative sensory testing (QST) can be used to precisely measure somatosensory function and pain sensitivity (Backonja et al., 2013; Rolke et al., 2006). Using QST, various types of noxious stimuli can be applied, including mechanical, thermal, and electrical stimuli, to determine the individual’s pain tolerance, thresholds, and other indicators of somatosensory function. Muddana et al. (2014) recently described cases of persistent breast and nipple pain without resolution using an attenuated QST assessment to identify differences in somatosensory functioning. The findings suggested a potential role for pain sensitization in the development of breast and nipple pain as has been identified in various other experimental and clinical pain outcomes (Diatchenko et al., 2005; Fillingim et al., 2005; Muddana, Asbill, Jerath, & Stuebe, 2018). While the emergence of painful conditions may be affected by development and environmental experiences, genetic factors contribute to individual differences in pain susceptibility across the lifespan. The present study was designed to assess for differences in somatosensory functioning (using QST) as well as determine the impact of pain-relevant genetic polymorphisms that may contribute to intolerable and/or ongoing breast and nipple pain susceptibility in breastfeeding women.

As the first study to explore the genetic factors contributing to ongoing breast and nipple pain, we chose to investigate relatively common pain-relevant genetic variants within catechol-o-methyltransferase (*COMT*) and the oxytocin receptor gene (*OXTR*) known to play a pivotal role in the neurohormonal cascade for breastfeeding. *COMT* and *OXTR* both have single-nucleotide polymorphisms (SNPs) with established associations with a variety of health behaviors (Baribeau et al., 2017; Diatchenko et al., 2005; Hu et al., 2018; Slane et al., 2014). The *COMT* SNPs rs4680 A>G (Val158Met), rs6269: A>G (promoter region), rs4633: C>T (His62His) and rs4818: C>G (Leu136Leu) have well-established associations with experimental pain sensitivity both in the normal healthy populations and in clinical populations with inflammatory, neuropathic, or postsurgical pain (Diatchenko et al., 2005; Hu et al., 2018). Given its role in catecholamine metabolism, it is perhaps unsurprising that *COMT* genotype is also associated with risk for depression, anxiety, and stress-reactivity (Antypa, Drago, & Serretti, 2013; Young et al., 2017). Relevant to the current study, *COMT* genotype has also been implicated as a genetic determinant of the length of the first stage of labor (Terkawi et al., 2012). Three haplotypes for *COMT*, composed of the combined genotypes for these four SNPs (rs4680 (A>G), rs6269 (A>G), rs4633 (C>T) and rs4818 (C>G) and strongly associated with pain sensitivity, have been identified as follows: low pain sensitivity (LPS) for G_C_G_G, average pain sensitivity (APS) for A_T_C_A, and high pain sensitivity (HPS) for A_C_C_G (Diatchenko et al., 2005). Oxytocin (OXT) has the potential to modulate pain through central and peripheral psychological and physiological mechanisms (Tracy, Georgiou-Karistianis, Gibson, & Giummarra, 2015; Xin, Bai, & Liu, 2017). The *OXTR* gene, encoding the receptor for OXT, contains two *SNPs*, rs2254298 (G>A) and rs53576 (G>A), known to affect parents sensitivity to their infants’ behavioral cues and is associated with depressive symptoms across the lifespan (Bakermans-Kranenburg & van Ijzendoorn, 2008; Saphire-Bernstein, Way, Kim, Sherman, & Taylor, 2011). Both *OXTR* rs53576 and *COMT* rs4633 SNPs contribute to increased length of early labor (Terkawi et al., 2012). In two large samples of early postpartum mothers, the *OXT* and *OXTR* polymorphisms were not associated with breastfeeding duration (Colodro-Conde et al., 2018), although a relationship with breast and nipple pain during breastfeeding was not explicitly assessed.

Clinically, the evaluation of breastfeeding pain is almost exclusively dependent on self-report measures by breastfeeding mothers. The most common evaluation used in breastfeeding studies to effectively capture changes in breast and nipple pain across time is the visual analogue scale (VAS) (Coca et al., 2018; McClellan et al., 2012). While the VAS is a validated global measure of pain severity or intensity, the VAS may not capture modality-specific differences in somatosensory aberrations that may contribute to pain sensitivity (i.e. pressure, mechanical, etc.) and may not accurately reflect the impact of pain (i.e. pain burden or pain interference) on function. A well-established objective measurement to detect somatosensory aberrations that may contribute to acute and chronic pain susceptibility is QST. QST has been used in chronic pelvic and low back pain conditions experienced in women (As-Sanie et al., 2013; Starkweather et al., 2016). In lactation, only one study attempted to use mechanical sensitivity testing using a toothpick on the breast (Muddana et al., 2018). However, no study has used a standardized mechanical QST protocol in women initiating breastfeeding to comprehensively evaluate breastfeeding pain.

## Research Question

Therefore, the aim of this secondary analysis was to examine associations among self-reported breast and nipple pain, experimental pain sensitivity using QST, and candidate pain sensitivity single-nucleotide polymorphisms [SNPs], (*COMT* rs6269, rs4633, rs4818, rs4680 and *OXTR* rs2254298, rs53576) in breastfeeding women.

## Methods

### Study Design

This study was a secondary analysis of selected pain-related data from a pilot randomized control trial which tested the feasibility and acceptability of a breastfeeding self-management (BSM) intervention among breastfeeding women (Lucas, Zhang, et al., 2019). The study was approved by the University of Connecticut Institutional Review Board in 2017, and registered with ClinicalTrials.gov (NCT03392675). Primary results of the study were used to determine effect sizes for future studies and have been reported elsewhere (Lucas, Zhang, et al., 2019).

### Setting

Women were recruited after delivery from one research-intensive tertiary medical center and one teaching community hospital in the northeast region of the United States.

### Sample

A convenience sample of 80 participants were approached and a total of 65 women were consented, five women withdrew before data collection, for a total of 60 women who completed data collection at entry, 1-, 2-, and 6 weeks. At 6 weeks, 56 women (26 BSM intervention, 30 control) were breastfeeding and their complete pain data set was used for this analysis. The pilot RCT was a feasibility study, with a sample size goal of 60 mothers which was large enough to report significant differences in average breastfeeding pain severity scores between the BSM and control groups (Erdfelder, Faul, & Buchner, 1996).

To be included, women 1) were 18 - 45 years of age; 2) English proficient; 3) delivered full term infant (38 – 42 weeks gestation); 4) planned to breastfeed, and 5) had daily access to a smartphone or computer. Women were excluded if they had a history of potential changes in pain sensorium, a significant mental health disorder (i.e. schizophrenia, bipolar disorder) or health condition(s) not associated with pregnancy (i.e. sickle cell anemia, HIV+, diabetes, history of seizures); or who delivered an infant with medical complications, congenital anomalies or ankyloglossia. In addition, participants were monitored throughout the study for infection, positioning challenges, infant ankyloglossia or torticollis, or abnormal nipple shape or size, and were withdrawn for any of these conditions.

### Breastfeeding Self-Management Intervention

Group assignment of women was based on a randomization schedule created by the study statistician. The BSM intervention was delivered via cloud based educational modules, bi-weekly nurse-lead texting, and weekly follow-up to women in their homes at 1-, 2-, and 6-weeks after discharge. A full description of the BSM intervention has been provided in a previous publication (Lucas, Bernier, et al., 2019). Members of the study team were blinded to group assignment.

### Measurements

Participants completed a demographic questionnaire, rated their breast and nipple pain intensity (0-100) using a visual analogue scale (Breivik et al., 2008), underwent QST using mechanical stimuli (cutaneous, vibration, pressure), and provided a buccal swab for genetic analysis. Both the BSM intervention and the control group received text/email at 1, 2, and 6 weeks, with a link to complete assessments for maternal report of breast and nipple pain severity.

### Data collection

The QST testing was collected in each participant’s hospital room before discharge. QST testing assessed peripheral and central pain sensitivity using a range of mechanical stimuli to measure Mechanical Detection Threshold, pain threshold, pain sensitivity, Windup Ratio (repeated mechanical pressure which captures temporal summation of pain), Vibration Detection Threshold, and Pain Pressure Threshold. The QST protocol and equipment followed standards of the German Neuropathic Pain Network (Rolke et al., 2006) with the exclusion of thermal testing to decrease participant burden.

Within 48 hours after delivery and before discharge, women were tested on the non-dominant arm for cutaneous, vibration, and pressure sensitivity and pressure pain thresholds. Women were asked to rate the severity of their experimental pain using a numerical rating scale (‘0’ as no pain to ‘10’ indicating ‘most intense pain imaginable’). Cutaneous Mechanical Pain Threshold and the Windup Ratio were assessed using von Frey fibers (Optihair2-Set, MarstockNervtest, Germany) which can exert a force between 0.25 and 512 mN upon bending. Dynamic Mechanical Allodynia was tested using a light brush stroke over 1 cm in length. The Vibration Detection Threshold used a Rydel–Seiffer tuning fork (64 Hz, 8/8 scale) and participants were asked to indicate when they no longer felt vibration. Lastly, Pressure Pain Thresholds were assessed by applying pressure manually at a range between 50 and 600 kPa using a Medoc algometer (Medoc Algomed, 2020). Participants held a button, which they pushed when they felt pain. The procedure took 10 minutes to complete.

Candidate pain sensitivity SNPs genotyping was conducted using buccal cell samples collected at baseline before discharge. Participants were instructed to rinse their mouth twice with water and then roll the sterile buccal brush firmly on the inside of the cheek. Samples were immediately transported to Dr. Young’s biobehavioral lab for processing and storage in a -80°C freezer for batch analyses. Genomic DNA was extracted from buccal cells using Gentra® Puregene® Buccal Cell Kit according to the manufacturer’s instructions (Qiagen, #158845). SNP genotyping was completed for seven SNPs; four SNPs within *COMT* and three within *OTXR* (Table 2) using Taqman SNP genotyping assays (VIC/FAM) and allelic discrimination analysis according to the manufacturer’s directions using an Applied Biosystems StepOne Plus PCR machine and ABI allelic discrimination software (ThermoFisher Scientific, Waltham, MA). Dr. Young verified a 95%≤ call rate for all SNPs included using 10uL samples and manufacturer’s running parameters.

### Data Analysis

Descriptive statistics were generated with mean and standard deviation for continuous variables, and frequency and proportion for discrete variables. Demographic characteristics, 7 QST measurements, and 6 SNPs for the 56 participants were included in the descriptive statistics table.

To evaluate the association between the SNP genotypes and pain sensitivity, we performed the Kruskal-Wallis one-way ANOVA on ranks to confirm if the distribution of a continuous QST was consistent in each genotype of a SNP. For the discrete QST measurement Dynamic Mechanical Allodynia, we used the Fisher’s Exact test instead.

Breast and nipple pain severity was the main outcome measurement considered in this secondary-analysis study. To visualize the effect of each SNP on breast and nipple pain severity, graphs with nonparametric smoothing curves were generated for pain severity over time by groups of alleles. Lucas et al. (2019) used a Linear Mixed Model (LMM) to evaluate the effect of the BSM intervention on reducing breastfeeding pain over 6 weeks. The model contained pain severity at week 1, 2, and 6 as the outcome, and baseline pain severity, week, BSM intervention group, and time by group interaction as covariates (Lucas, Zhang, et al., 2019). In this secondary-analysis, a similar LMM model with adjusted demographic variables was considered. We put each QST and SNP into the model alternatively, and conducted the Likelihood Ratio Test (LRT) to verify the significant effect of each QST and SNP on reducing breastfeeding and nipple pain over time. We coded the number of minor alleles (0-2) as the covariate of SNP in order to minimize the degrees of freedom due to the small sample size. All the analyses were performed with the statistical software R 3.6.2.

## Results

### Demographics and Allelelic Frequency

There was no significant difference in demographic characteristics (age, race or ethnicity, delivery or parity) between the two intervention groups (see Table 1). The mean age of the participants was 30.38 years old (SD = 4.86 years). Participants were mainly Caucasian (76.8%), non-Hispanic or Latino (76.8%) and 80.4% had a vaginal delivery. Fewer than half (46.4%) were breastfeeding naive. Means and standard deviations of QST measures (*see* Table 1) were reported except for the Dynamic Mechanical Allodynia as the majority of patients scored a 0 (83.9%), so we treated it as a categorical variable (=0 or 0<). Allelic frequencies and relative proportions for all SNPs were reported in Table 1. Sample *OXTR* rs2254298 and rs53576 and *COMT* rs6269 and rs4818 minor allele frequency were similar to global frequency but *COMT* rs4633 (Sample 0.51, Global 0.37), and rs4680 (Sample 0.49, Global 0.37) minor allele frequencies were higher than global values.

**Table 1.**
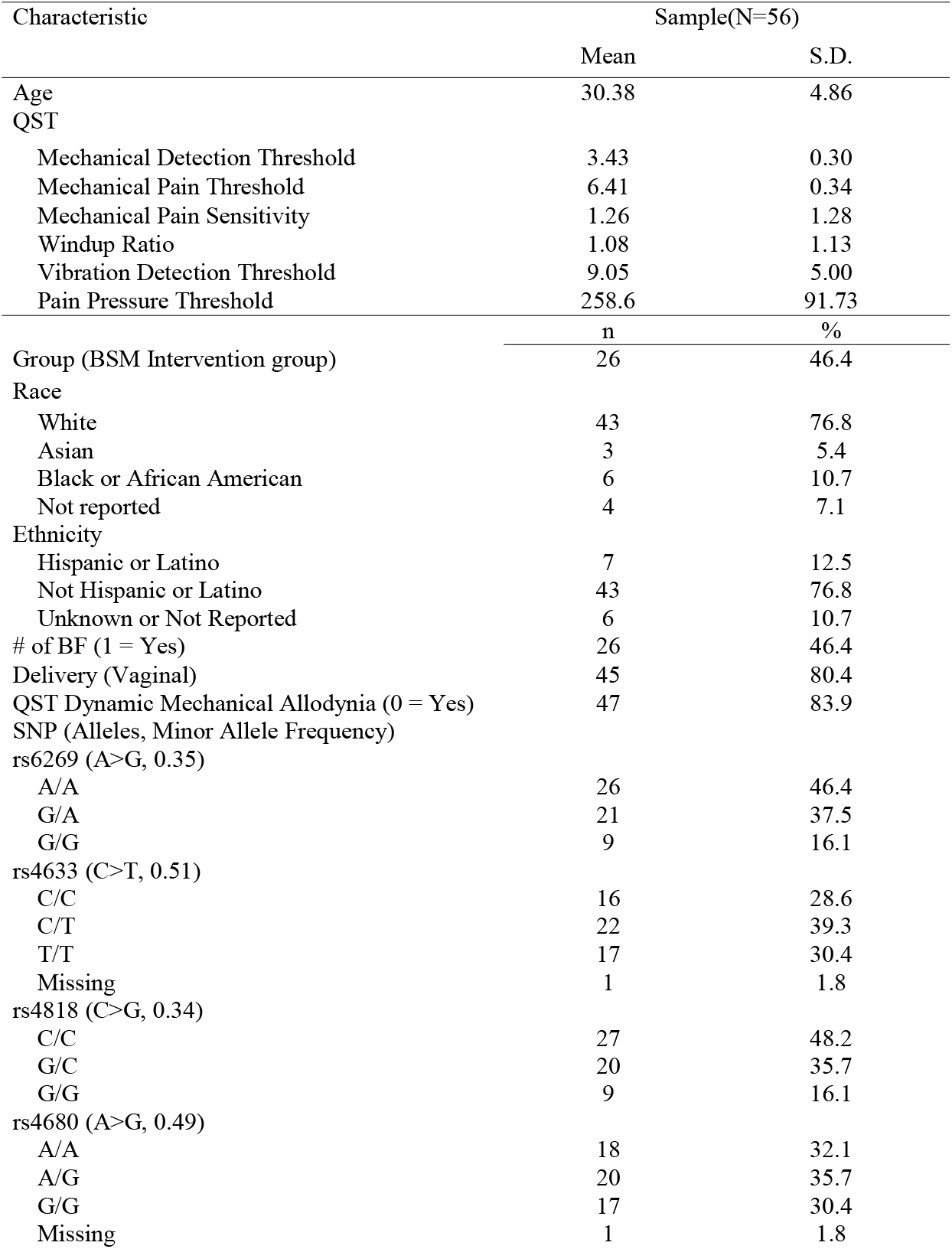
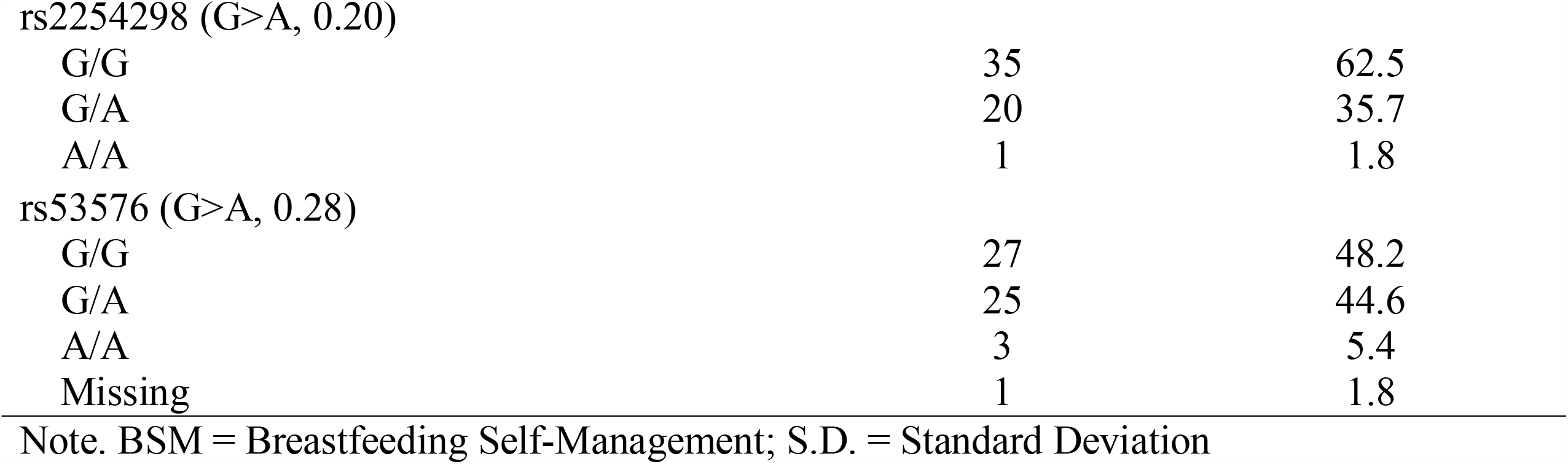
Demographic Characteristics

### Pain Severity Over Time vs SNPs

Pain severity was plotted over time for each SNP genotype using the Local Polynomial Regression Fitting method provided by ‘geom_smooth’ function in ‘ggplot2’ package. Only rs53576 genotype was significantly associated with pain severity reports over time. Participants who were heterozygotes (A/G; green curve) and homozygous (G/G; blue curve) reported similar baseline pain severity, but the G/G group reported significantly lower pain severity compared to the A/G group at weeks 1, 2, and 6.

Table 2 presents the findings of the LRT result for each SNP, respectively. Only the minor allele of rs53576 had a significant effect on reducing breast and nipple pain severity by time (p = 0.036). The estimated coefficient of the number of minor allele of rs53576 is 8.18, which can be interpreted that, on average, the women experienced an 8.18 increase in breastfeeding and nipple pain scores over time with the addition of one minor allele A of rs53576. The time by number of allele interaction effect of rs53576 was also checked, but it was not significant. No significant effect was found for the other SNPs.

**Table 2.**
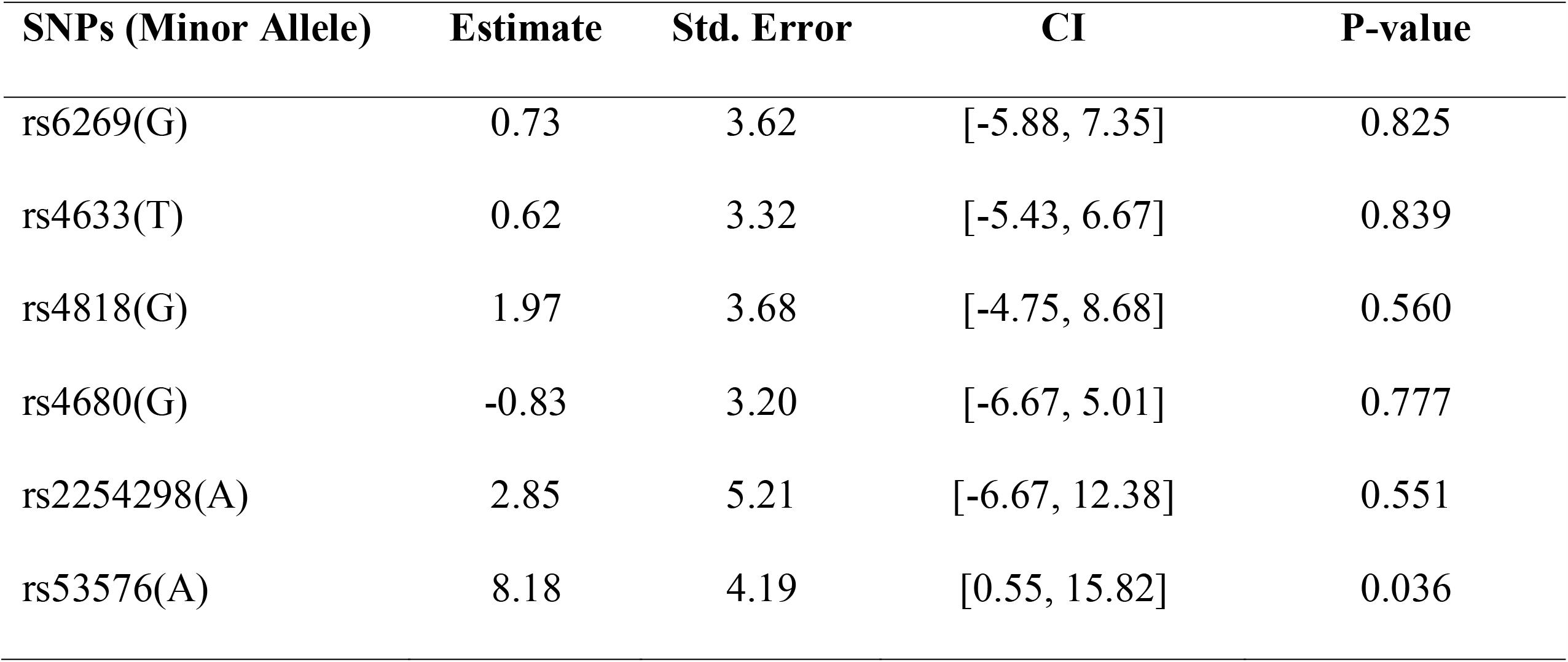
Estimated Coefficients, and LRT Result of Each SNP

### Pain Severity and Somatosensory Function (QST)

Table 3 presents the findings of LRT for each QST variable, respectively. Significant effects were found for two QSTs standardized tests, Mechanical Detection Threshold (sensation) (p = 0.036) and Windup Ratio (repeated mechanical stimulation) (p = 0.033). The estimated coefficient of the Mechanical Detection Threshold and the Windup Ratio were 16.51 and 4.82, respectively, indicating that participants experienced 16.51-fold higher breast and nipple pain scores over time if they were 1-unit more sensitive in the measurement of Mechanical Detection Threshold. Women also experience 4.82-fold higher breast and nipple pain severity scores over time if they were 1-unit more pain sensitive in the measurement of Windup Ratio. There was no significant interaction effect with time for these two QSTs. No significant effect was found for the other QST tests.

**Table 3.**
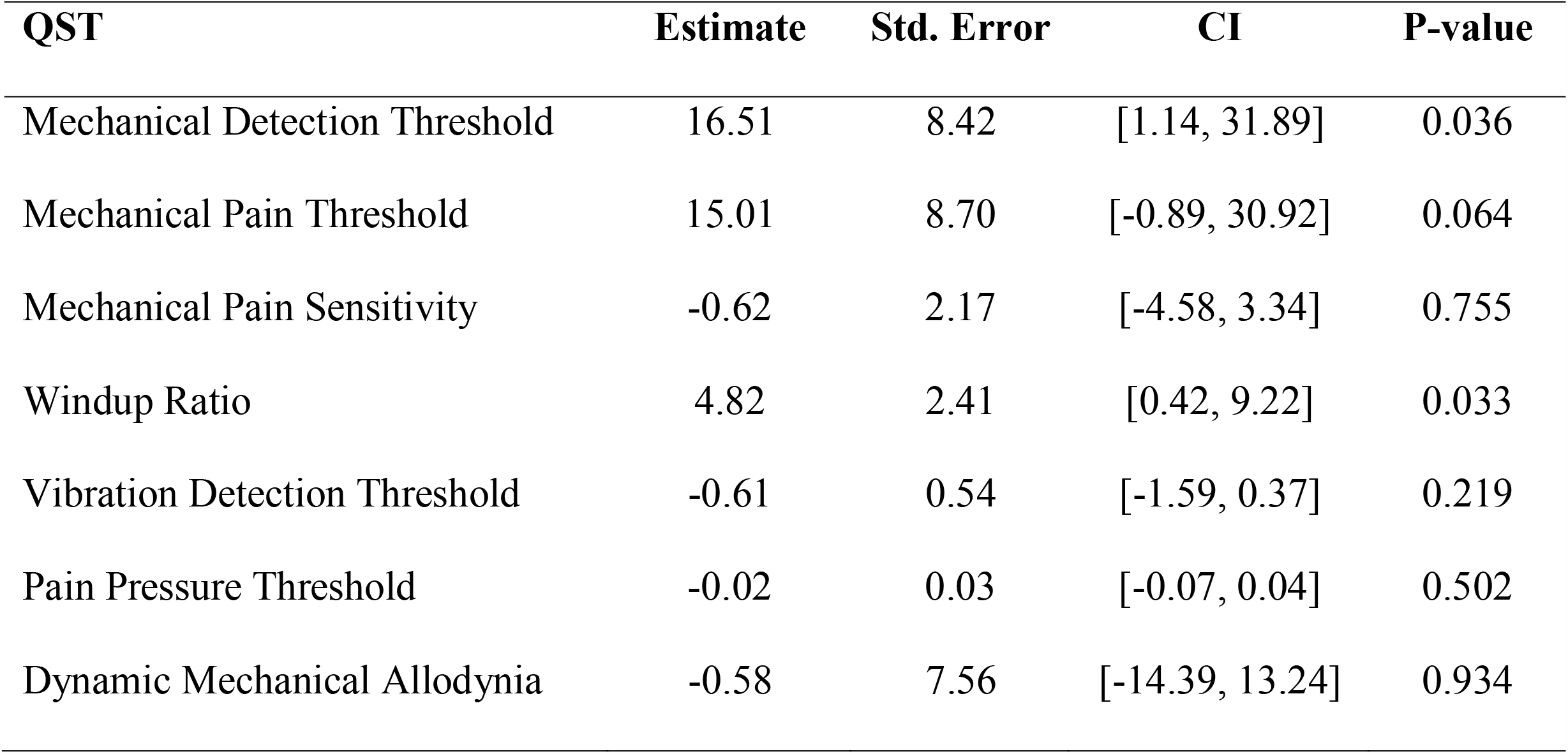
Estimated Coefficients, and LRT Result of Each QST

### SNPs associations with somatosensory function (QST)

Due to the non-parametric distribution of QST measurements, Kruskal-Wallis one-way ANOVA was performed for the association between QSTs and SNPs. For the discrete QST measurement Dynamic Mechanical Allodynia, we performed Fisher’s Exact test to check its association with each SNP. Only Dynamic Mechanical Allodynia and rs2254298 had a significant association (p = 0.03) (Table 4). Thirty-two women with G/G and 15 women with A/G had Dynamic Mechanical Allodynia value 0, and 3 women with G/G, 5 women with A/G and 1 woman with A/A had Dynamic Mechanical Allodynia value greater than 0. It shows that the number of minor alleles of rs2254298 increased the likelihood of a non-zero Dynamic Mechanical Allodynia value. No other significant associations were found between QST measures and SNP genotypes.

**Table 4.**
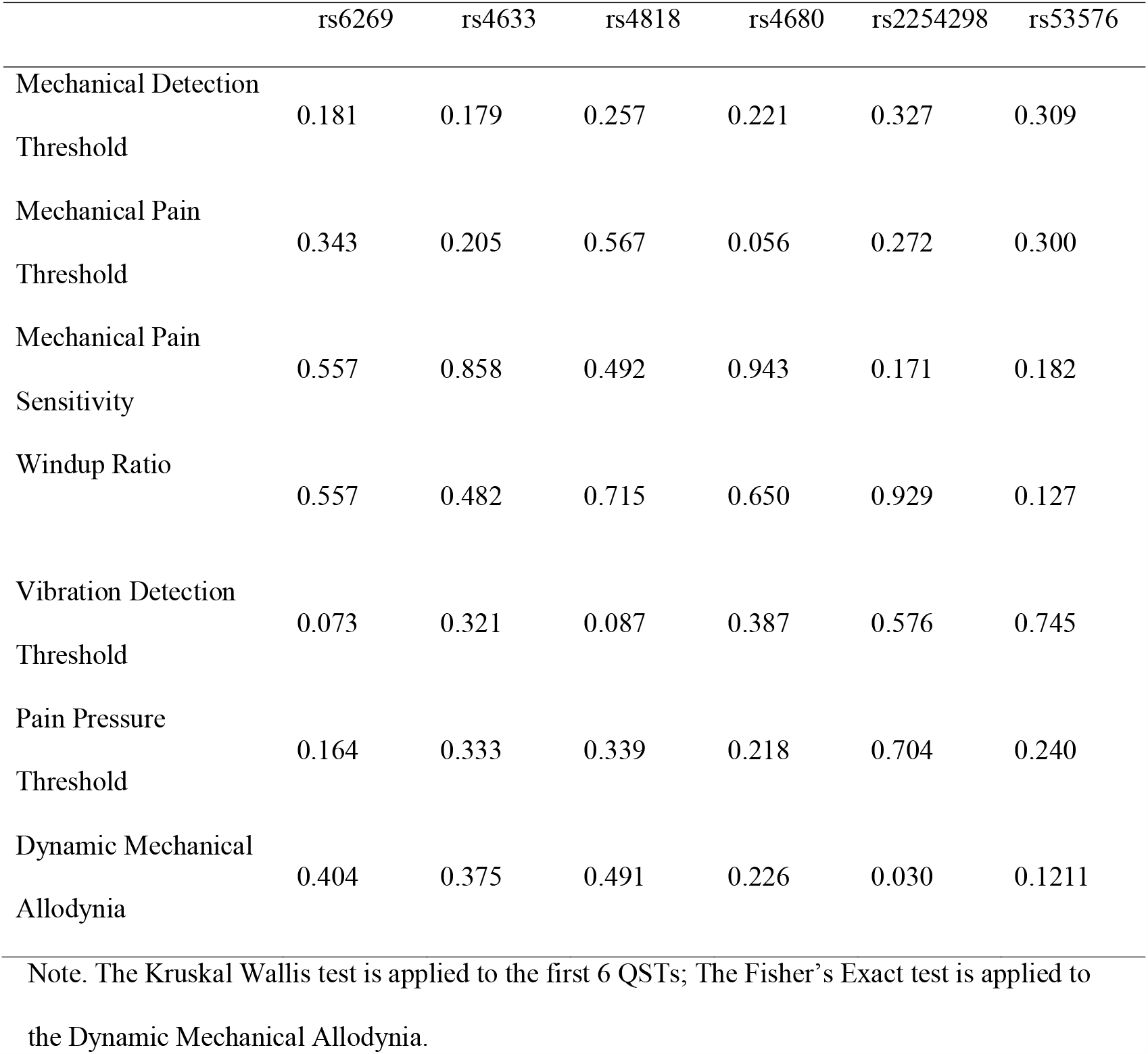
P-values of hypothesis testing QST vs SNP.

## Discussion

The present study explored associations among self-reported breast and nipple pain, somatosensory function (QST) and pain sensitivity polymorphisms in women who were initiating breastfeeding. In our pilot sample, only women with the minor allele *OXTR* rs53576 reported significantly higher breast and nipple pain severity over time, with 8.18-fold higher pain scores compared to women with the major allele. In addition, the QST standardized test, Mechanical Detection Threshold and Windup Ratio (repeated mechanical stimulation) were predictive of greater self-report pain severity. For every 1-unit sensitivity to the Mechanical Detection Threshold, women experienced 16.51 more breast and nipple pain scores and for the Windup Ratio, women reported 4.82 higher breast and nipple scores. Lastly, women who had one or more minor alleles at *OXTR* rs2254298 were more likely to experience hypersensitivity to brush stimulation in the Dynamic Mechanical Allodynia test.

OXT is a significant neurohormone throughout the perinatal period and breastfeeding initiation (Augustine, Seymour, Campbell, Grattan, & Brown, 2018). Oxytocin signaling through *OXTR* genotype in the peripheral and central nervous system plays a critical role in sensory processing and pain perception (Xin et al., 2017). Although no prior studies have directly linked *OXT* and/or *OXTR* genotype to breastfeeding outcomes, our study suggests that *OXTR* genotype has a moderating effect on breast and nipple pain and thereby may contribute to breastfeeding success and duration. Women carrying 1 or more of the minor alleles at *OXTR* rs53576 had a significantly higher pain score and reported earlier sensitivity to mechanical pressure and repeated sensations for the Windup Ratio test. For women with *OXTR* rs53576 minor alleles, the increased pain scores at 1 and 2 weeks after delivery may be a response to the 8 to 12 times a day breastfeeding sessions in which women potentially experience repeated pressure and somatosensory painful simulation until the lactation tissue becomes desensitized or infant positioning has been corrected (Lucas & McGrath, 2016). Women with *OXTR* rs2254298 regardless of heterozygous or homozygous major and minor alleles were more likely to experience allodynia.

In our study, we anticipated that differences in self-report pain scores would be associated with *COMT* gene minor alleles. Although, the *COMT* rs4633, and rs4680 alleles were higher in our sample than global frequency, there was no significant relationship between women’s pain severity scores and the *COMT* gene. Future studies will need to extend the present findings to explore the impact of genotype on breast and nipple pain in breastfeeding beyond the acute pain of breastfeeding compared to ongoing chronic pain at 3 and 6 months.

### Clinical Implications

In the present study, QST provided a simple and non-invasive technique by which to identify women at risk of high levels of pain during breastfeeding. This may serve as a method for nurses to identify, counsel and support women at risk as they begin breastfeeding with the goal of promoting exclusive breastfeeding throughout the 6-month period.

Women can experience ongoing pain during breastfeeding due to mechanical pain of a poor infant latch or positioning, or shearing pain from the wrong size of flange of their pump (Lucas & McGrath, 2016). In alignment with these prior findings, infant-related issues were indicated as the reason for discontinuation of breastfeeding in all four women who stopped breastfeeding during the study, three women from the control group due to infant ankyloglossia or tongue-tie, and one woman from the intervention group due to poor infant latch. All four women reported ongoing elevated pain scores and transitioned to pumping breast milk.

Women reporting breast and nipple pain should first be assessed for other potential pain triggers including infection (e.g., *Candida*, bacterial mastitis) or underlying medical conditions, (e.g. psoriasis or Reynaud’s syndrome) (Berens et al., 2016; Lucas & McGrath, 2016). If treatment of these conditions does not resolve pain, clinicians should consider the presence of allodynia or other neuropathic pain syndrome, and prescribe medications recommended by American Breastfeeding Medicine protocol (Berens et al., 2016). In addition, as emotional regulation of anxiety and depression contribute to women’s pain sensorium, clinicians should consider referring women for counseling or peer support groups, encouraging non-pharmacological interventions of creative imagery and massage therapy or pharmacological intervention of a breastfeeding safe antidepressant (Sriraman, Melvin, & Meltzer-Brody, 2015; Subnis, Starkweather, & Menzies, 2016). Even with clinical interventions to manage ongoing pain, many women cease breastfeeding due to returning to work or a lack of social support (Center for Disease Control, 2018). These environmental and social factors may differentially affect those with increased pain susceptibility, further adding to the conflict women experience regarding ceasing breastfeeding and their role as a mother providing optimal nutrition to their infant (Jackson et al., 2019).

The U.S. Preventative Task Force, recognizes that women experience pain of greater severity and frequency than men (US Preventive Task Force, 2016). As breastfeeding is considered a” normal” health behavior, breastfeeding pain has not been explored as a “pain” condition has not been explored. If a woman has increased susceptibility for breast and nipple pain as a result of her genotype, and she experiences ongoing pain, this could lower her mechanical pain threshold and put her at-risk for chronic pain in the future. In addition, for women who present with a non-breastfeeding, but chronic pain condition, a breastfeeding history with targeted questions about breastfeeding being an ongoing pain experiences should be explored in the individual’s pain history.

### Limitations

This is a secondary analysis of a pilot study evaluating pain related to genetic risk. As a small study, this has a risk to produce false positive and negative results which needs a larger study to validate the results. In addition, we used the visual analogue scale (0 – 100) which is reliable to measure acute pain, before and after an intervention but clinical is not as sensitive to small differences in pain scores (Breivik et al., 2008). Also due to the small sample size, group was not factored into the results and the report of pain might be higher without the moderation of the breastfeeding pain intervention.

A confounding factor of women’s self-report of pain, is the standard of care during delivery, many women receive synthetic oxytocin during and after delivery. Women receiving synthetic oxytocin during labor experience a downstream effect of decreased endogenous oxytocin affecting breast milk supply for several weeks (Cadwell & Brimdyr, 2017). We hypothesize that some women may present with allodynia as a result of labor-related exogenous oxytocin due to the blockage of *OXTR* receptor sites, and barriers to endogenous oxytocin. In our pilot study, 75% of women received oxytocin augmentation our data suggest that their threshold of mechanical sensitivity, pain, and allodynia could be affected by the action of exogenous oxytocin. We also hypothesize, that women with minor *OXTR* rs53576 and rs2254298 alleles lack the moderating effect of oxytocin and with the frequent number of feedings have greater pain. In future studies, we will collect the total amount of exogenous oxytocin mothers receive during labor to see if there is an effect on the endogenous expression of oxytocin related to *OXTR* alleles.

## Conclusion

Women with ongoing breast and nipple pain after breastfeeding initiation suggests a phenotype of genetic risk for BF pain to identify women at high risk of pain-associated BF cessation. In addition, the novel application of somatosensory testing using QST for sensitivity and pain sensation thresholds in the postpartum period, identified women who reported higher breast and nipple pain during the first six weeks after delivery. Together these findings provide an objective assessment of women at-risk for pain during the postpartum period.

**Figure 1.**
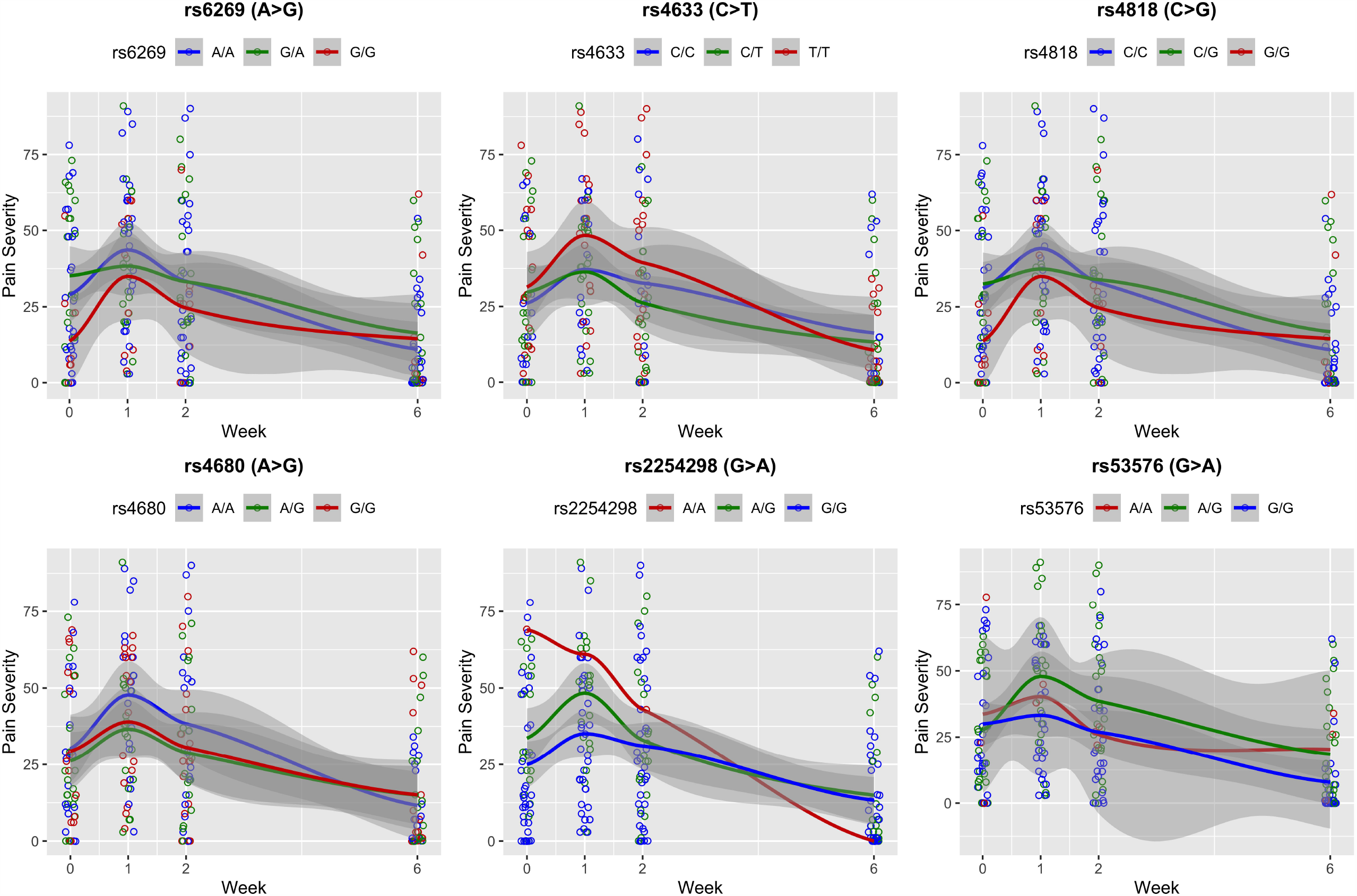
The red curve in each plot represents the trend of mean pain severity for the homozygous minor (rs6269, rs4818, rs4680, GG; rs4633, TT; rs2254298, rs53576, AA); the green curve represents the heterozygous minor alleles (rs6269, G/A; rs4818 C/G; rs4680 A/G; rs4633, C/T; rs2254298, rs53576, A/G), and the blue curve was for the homozygous major (rs6269, rs4680, A/A; rs4818, rs4633, C/C; rs2254298, rs53576, A/A). The grey shade represented the confidence interval of the trend. Note that there was only one A/A of rs2254298 in the sample, so the red curve in the rs2254298 graph represents the pain severity trend for that single individual.

## Data Availability

Deidentified symptom data (including the data dictionary) will be provided to interested scientific investigators for the purpose of secondary research analysis by contacting the corresponding author and sending the investigators curriculum vitae and research questions. The study protocol and statistical analysis plan is available in our previous publication.

https://cdrns.nih.gov/

## Acknowledgement

Research reported in this publication was supported by the National Institute of Nursing Research of the National Institutes of Health (NIH-NINR), Grant number: NIH-NINR P20NR016605. The content is solely the responsibility of the authors and does not necessarily represent the official views of the National Institutes of Health. The authors thank Dr. Deborah McDonald, RN, PhD, for her support and advice.

